# Assessment Of Household Food Security and Nutritional Status of Mother-Child Pair in Selected Local Government Areas in Ondo State

**DOI:** 10.64898/2026.04.29.26352102

**Authors:** Gbayisemore Erisan Eunice, Kayode Ajayi, Ibidayo Adefolake Alebiosu, Abolaji Moses Ogunetimoju

**Affiliations:** Department of Human Nutrition and Dietetics, Afe Babalola University, Ado Ekiti, Nigeria; Department of Human Nutrition and Dietetics Afe Babalola University, Ado Ekiti, Nigeria; Department of Statistics, Obafemi Awolowo University, Osun State, Nigeria

**Keywords:** Food Security, Malnutrition, Stunting, Wasting, BMI, Infant Feeding Practices, Ondo State, Nigeria

## Abstract

**Introduction:** Food insecurity and mixed patterns of malnutrition coexist in rural Nigeria despite the country’s agricultural potential. There is a lack of precise data regarding the relationship between household food security and nutritional status of the mother-child pair in southwestern Nigeria. This study examined household food security and mother-child nutritional status in Irele and Okitipupa Local Government Areas (LGAs) of Ondo State.

**Methods and Analysis:** A descriptive cross-sectional community-based survey was carried out with 358 mother-child pairs (children 6-59 months). The Household Food Insecurity Access Scale (HFIAS) was used to measure household food insecurity. Body Mass Index (BMI) was used to assess mothers’ and WHO Anthro Z-scores to assess children’s nutritional status. Descriptive statistics and Chi-square tests (p < 0.05) were used to examine the data.

**Results:** 93.3% of households were food insecure. A striking double burden of malnutrition was observed: 58.4% of mothers were overweight or obese, and child malnutrition was widespread, with 39.3% stunting, 29.1% wasting and 42.1% underweight. Breastfeeding duration (p = 0.008) and introduction of complementary feeding (p = 0.032) were significant predictors of child wasting. Interestingly, maternal education and income were not significant predictors of child undernutrition (p > 0.05), suggesting that environmental and behavioral influences take precedence over individual socioeconomic status in these communities

**Conclusion:** The simultaneous presence of severe child undernutrition and maternal overnutrition in rural Ondo State suggests a public health crisis in a state undergoing nutrition transition towards energy-dense, low-nutrient foods. These results suggest that national approaches are inadequate. We need interventions that focus on decentralized, LGA-level policies that integrate food security programs with education on Infant and Young Child Feeding (IYCF).

**What is already known on this topic:** Household food insecurity is a major driver of malnutrition among mothers and children under five in Nigeria’s rural communities. Evidence from southwestern Nigeria shows that over 88% of rural households are food insecure, with women and young children disproportionately affected. However, localized data from specific LGAs in Ondo State—particularly examining the mother-child dyad—remains scarce.

**What this study adds:** This study provides the first localized evidence from Irele and Okitipupa LGAs, Ondo State, documenting a 93.3% household food insecurity rate and a dual burden of malnutrition (39.3% child stunting and 58.4% maternal overweight/obesity coexisting in the same communities). It demonstrates that IYCF practices—specifically breastfeeding duration and timing of complementary food introduction—are significant determinants of child wasting, and highlights the limitations of maternal socioeconomic variables alone as predictors of child nutritional outcomes.

**How this study might affect research, practice or policy:** For research, this study establishes a dyadic methodological framework applicable to other Nigerian states. For practice, it underscores that nutrition education on IYCF practices must accompany food security programs. For policy, the findings call for decentralized, LGA-specific strategies addressing both rural food insecurity and the emerging nutrition transition—moving beyond one-size-fits-all national approaches to combat simultaneous undernutrition and overnutrition within the same households.

## 1.0 INTRODUCTION

Food security and adequate nutrition are central to global health and development, particularly in low- and middle-income countries. The United Nations Sustainable Development Goal 2 (Zero Hunger) seeks to end all forms of hunger and malnutrition by 2030, emphasizing equitable access to sufficient, safe, and nutritious food (Atukunda et al., 2021).

Despite Nigeria’s substantial agricultural capacity, household food insecurity remains widespread, especially in rural regions where poverty, infrastructural deficits, and low maternal education exacerbate vulnerability (Adeyanju & Fadupin, 2024). Food insecurity extends beyond the mere absence of food to encompass restricted access, poor dietary diversity, and inadequate utilization of available food for optimal nutritional outcomes (Olamide, 2024).

Recent studies across Nigeria have reported alarmingly high rates of food insecurity among rural households, with over 88% of families in some regions categorized as food insecure, ranging from mild to severe hunger (Adeyanju & Fadupin, 2024). These conditions disproportionately affect women and children, particularly those under five years of age who are at a critical stage of growth and development.

Malnutrition in children aged 6–59 months is strongly associated with food insecurity, manifesting in high prevalence rates of stunting, wasting, and underweight—conditions linked to increased morbidity and mortality (Olodu et al., 2022). Mothers, who serve as primary caregivers and food providers in Nigerian households, face additional challenges such as inadequate income, limited nutrition education, and insufficient control over food-related decisions.

A study in Ibadan revealed that maternal roles in food safety and hygiene are frequently constrained by limited access to clean water, shared kitchen spaces, and poor household infrastructure (Atoloye et al., 2024). Ondo State, despite being a predominantly agricultural region, lacks adequate localized data on the interrelationship between household food security and child nutritional status. In particular, rural Local Government Areas (LGAs) such as Irele and Okitipupa remain underexplored, raising critical questions about access, equity, and utilization—dimensions often overlooked in rural nutrition planning. This contradiction between national food availability and household food insecurity underscores the complex and multifaceted nature of hunger in Nigeria and highlights the need for localized evidence to inform effective interventions.

Therefore, this study aims to assess household food security and the nutritional status of mothers and their children aged 6–59 months in Irele and Okitipupa LGAs of Ondo State. The study is guided by the null hypothesis that there is no significant relationship between household food security and the nutritional status of mothers and children aged 6–59 months, and the alternative hypothesis that such a significant relationship does exist. The significance of this study lies in its potential to provide empirical evidence on the intersection of food security and nutrition in rural Ondo State, thereby contributing to the understanding of how household-level vulnerabilities shape maternal and child health outcomes.

By identifying the extent to which food insecurity influences nutritional status, infant and young child feeding practices, diet quality, and sanitation, the findings will inform policymakers, health practitioners, and development agencies in designing context-specific interventions. Ultimately, the study will help bridge the gap between agricultural production and equitable nutritional outcomes, offering insights that are critical for achieving Zero Hunger and improving the health and wellbeing of vulnerable populations in Nigeria.

## 2.0 METHODOLOGY

A descriptive cross-sectional community-based survey design was employed to provide a snapshot of the study population at a single point in time. This design was appropriate for assessing patterns, relationships, and variations in household food security and nutritional outcomes across different socioeconomic groups within rural communities.

The study was conducted in Irele and Okitipupa Local Government Areas (LGAs) of Ondo State, Nigeria, between June and July 2025. These LGAs are predominantly rural and semi-urban, with most residents engaged in small-scale farming, fishing, and informal trading. The combined population of both areas served as the sampling frame. These areas were purposively selected because of documented challenges related to food availability, child nutrition, and limited prior localized research. The study population comprised mothers aged 15–49 years and their children aged 6–59 months who had resided in the selected communities for at least six months. Children under five are particularly vulnerable to malnutrition, and mothers play a central role in their feeding and care. Mothers or children who were severely ill were excluded from the study.

To ensure the results are accurate and reliable, we must calculate the right number of people to include in the study. A total of 300 mother-child pairs will be selected using Cochran’s formula with 95% confidence level and 5% margin of error. The sample size will be determined using the Cochran formula for estimating sample size in prevalence studies:

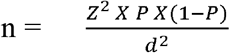

Where:

**Where:**

➢ n= sample size
➢ Z = standard score for 95% confidence = 1.96
➢ p = estimated proportion of households that are food insecure = 0.5 (used when the actual figure is unknown)
➢ d= margin of error = 0.05

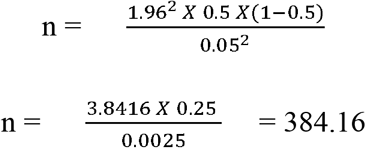

A multistage sampling method was employed.

In Stage 1, Irele and Okitipupa LGAs were purposively selected.

In Stage 2, three wards per LGA were randomly selected by lottery.

In Stage 3, households with children aged 6–59 months were systematically sampled within selected wards, with one eligible adult per household randomly selected.

Data were collected through face-to-face interviews using structured questionnaires covering socio-demographic and economic characteristics, household food insecurity (HFIAS), infant and young child feeding practices, dietary patterns, and sanitation and hygiene practices. Nutritional status was assessed via anthropometric measurements (weight, height/length) used to calculate BMI for mothers and WHO Z-scores for children using WHO Anthro software.

Questionnaires were administered in English or Yoruba by trained field assistants. Content and face validity were established through expert review, while reliability was assessed using a test-retest method among 20 participants, yielding an acceptable correlation coefficient. Data analysis was conducted using SPSS. Descriptive statistics (means, standard deviations, frequencies, and percentages) were used to summarize variables. Inferential statistics (Chi-square tests) were applied to test associations at a significance level of p < 0.05. Child nutritional indicators were classified per WHO thresholds: stunting (HAZ < −2 SD), wasting (WHZ < −2 SD), underweight (WAZ < −2 SD), and BMI-for-age Z-score (BAZ).

Ethical approval was obtained from the Health Research Ethics Committee (HREC) of the Ministry of Health, Ekiti State, Nigeria, in accordance with the Declaration of Helsinki. Informed consent was obtained from all participants, and confidentiality and anonymity were strictly maintained throughout. Patients or members of the public were not involved in the design, conduct, reporting, or dissemination plans of this research. Participants provided primary data through structured surveys and nutritional assessments but did not serve as active collaborators in data analysis or manuscript preparation. Findings will be disseminated to relevant health authorities in Ondo State and shared with participating communities through appropriate institutional channels.

## 3.0 RESULTS

A total of 358 mother–child pairs were included in the final analysis. The key findings across all study objectives are presented below. Household food insecurity was nearly universal in the study communities. The majority of households (93.3%) were food insecure, with most classified as mildly food insecure (88.8%). This reflects severe structural limitations in food access and stability among rural households in Irele and Okitipupa LGAs. The findings highlight the persistence of food insecurity despite the agricultural potential of the region, underscoring the gap between food availability at the national level and household-level access and utilization.

Table 1 presents the nutritional status of the 358 mothers by Body Mass Index (BMI). The results show a dual burden of malnutrition among mothers. While 5.9% were underweight, the majority were overweight (19.6%) or obese (38.8%), with only 35.8% in the normal BMI range. This indicates that overnutrition is more prevalent than undernutrition, reflecting the nutrition transition where energy-dense but nutrient-poor diets contribute to rising overweight and obesity even in food-insecure households.

**Table 1:** Nutritional Status of Mothers (BMI) (n = 358)

| *BMI Category* | *BMI Cut-off* | *Frequency* | *Percentage (%)* |
| --- | --- | --- | --- |
| *Underweight* | < 18.5 kg/m<sup>2</sup> | 21 | 5.9 |
| *Normal weight* | 18.5–24.9 kg/m<sup>2</sup> | 128 | 35.8 |
| *Overweight* | 25.0–29.9 kg/m<sup>2</sup> | 70 | 19.6 |
| *Obese* | ≥ 30.0 kg/m<sup>2</sup> | 139 | 38.8 |

**Source:** Field survey, 2025

Table 2 presents the nutritional assessment of under-five children reveals a complex picture of both undernutrition and overnutrition. Wasting affected 29.1% of children (15.8% severely and 13.3% moderately), which is nearly double the WHO emergency threshold of 15%, indicating a critical public health concern. Stunting prevalence was 39.3% (28.2% severely and 11.1% moderately), reflecting chronic growth failure and long-term nutritional deprivation. Underweight was also widespread, with 42.1% of children falling below normal weight-for-age, including 28.4% severely underweight. At the same time, overweight and obesity were present: 14.4% of children were overweight by WAZ, and 25.1% were overweight or obese by BMI-for-age Z-scores (BAZ). This coexistence of high levels of wasting, stunting, and underweight alongside overweight and obesity demonstrates a dual burden of malnutrition within the same population. It suggests that while many children suffer from inadequate dietary intake and poor growth, others are exposed to energy-dense but nutrient-poor diets that predispose them to overnutrition.

**Table 2:** Nutritional Status of Under-Five Children by WHO Anthro Z-Score (n = 358)

| *Nutrition Status Classification* | *Percentage (%)* | *Frequency* |
| --- | --- | --- |
| *WASTING (WHZ)* |  |  |
| Severe wasting | 15.8 | 45 |
| Moderate wasting | 13.3 | 38 |
| Normal | 70.9 | 202 |
| *STUNTING (HAZ)* |  |  |
| Severely stunted | 28.2 | 61 |
| Moderately stunted | 11.1 | 24 |
| Normal | 60.6 | 131 |
| *UNDERWEIGHT (WAZ)* |  |  |
| Severely underweight | 28.4 | 77 |
| Underweight | 13.7 | 37 |
| Normal | 43.5 | 118 |
| *Overweight* | 14.4 | 39 |
| *BAZ* |  |  |
| Severely thin | 15.6 | 48 |
| Thin | 16.3 | 50 |
| Normal | 43.0 | 132 |
| *Overweight* | 4.9 | 15 |
| *Obese* | 20.2 | 62 |

**Source:** Field survey, 2025

Overall, the findings highlight a community facing both acute and chronic undernutrition, coupled with rising overnutrition, underscoring the urgent need for integrated interventions that address food insecurity, diet quality, and child feeding practices simultaneously.

Tables 3 shows that certain infant feeding practices were significantly associated with child wasting. Breastfeeding duration had a strong relationship (p = 0.008): children who were severely wasted were more likely to have mothers who breastfed for shorter periods (< 6 months), while those breastfed for one to two years had lower rates of wasting and higher proportions of normal nutritional status. Similarly, the timing of family food introduction was significant (p = 0.032). Early introduction of family foods (< 6 months) was linked to higher rates of severe wasting, whereas introduction at six months was associated with better outcomes. Exclusive breastfeeding, however, showed no significant association with wasting (p = 0.758). Both wasted and normal children had similar proportions of exclusive breastfeeding, suggesting that duration and timing of complementary feeding may play a more critical role than exclusivity alone in this population. Overall, the findings highlight that shorter breastfeeding duration and early family food introduction are risk factors for wasting, while adherence to recommended practices (breastfeeding up to 2 years and introducing family foods at 6 months) is protective.

**Table 3:** Infant Feeding Practices vs Wasting (WHZ)

| *Variable* | *Severe Wasting n (%)* | *Moderate Wasting n (%)* | *Normal n (%)* | *P-value* |
| --- | --- | --- | --- | --- |
| *Breastfeeding Duration* |  |  |  | 0.008 |
| $< 6$ months | 3 (6.7) | 11 (28.9) | 17 (8.4) | |
| 6 months | 7 (15.6) | 4 (10.5) | 22 (10.9) |  |
| 1 year | 15 (33.3) | 14 (36.8) | 83 (41.1) |  |
| 2 years | 20 (44.4) | 9 (23.7) | 80 (39.6) |  |
| *Family Food Introduction* |  |  |  | 0.032 |
| < 6 months | 15 (33.3) | 8 (21.1) | 85 (42.1) |  |
| 6 months | 25 (55.6) | 19 (50.0) | 89 (44.1) |  |
| 1 year | 5 (11.1) | 11 (28.9) | 28 (13.9) |  |
| *Exclusive Breastfeeding* |  |  |  | 0.758 |
| Yes | 24 (53.3) | 21 (55.3) | 100 (49.5) |  |
| No | 21 (46.7) | 17 (44.7) | 102 (50.5) |  |

**Source:** Field survey, 2025

Table 4 clearly reveals that, across all four anthropometric indicators—wasting, stunting, underweight, and BMI-for-age Z-scores—Chi-square analysis revealed no statistically significant associations with monthly income, education level, or occupation. This suggests that within this relatively homogeneous low-income population, malnutrition is cross-cutting and not differentiated by individual socioeconomic status. The persistence of both undernutrition and overnutrition points instead to broader structural and environmental determinants, such as community-level food insecurity, poor dietary diversity, and inadequate health services, which affect all households regardless of socioeconomic differences.

**Table 4:** Socioeconomic Status and Child Nutritional Outcomes.

| *Socioeconomic Variable* | *Wasting (p = 0.605)* | *Stunting (p = 0.168)* | *Underweight (p = 0.964)* | *BAZ (p = 0.182)* |
| --- | --- | --- | --- | --- |
| *Monthly Income* | No significant association | No significant association | No significant association | No significant association |
| *Education Level* | No significant association | No significant association | No significant association | No significant association |
| *Occupation* | No significant association | No significant association | No significant association | No significant association |

**Source:** Field survey, 2025

From table 5, no statistically significant associations were found between sanitation and hygiene practices—including feeding utensil type, utensil storage, water source, water treatment, and toilet facility—and child nutritional outcomes (p-values ranged from 0.173 to 0.865). This suggests that malnutrition in the study communities is not differentiated by household-level sanitation practices. However, a notable finding was that 66.8% of mothers reported not treating domestic water, a pattern consistent across all nutritional status groups. This indicates a pervasive community-level risk factor that may contribute to poor health and nutrition outcomes broadly, rather than serving as a distinguishing variable between children of different nutritional statuses.

**Table 5:** Sanitation, Hygiene, and Child Nutritional Status.

| *Practice* | *Underweight ($p = 0.388\text{--}0.710$)* | *BAZ ($p = 0.173\text{--}0.865$)* |
| --- | --- | --- |
| *Feeding utensil type* | No significant association | No significant association |
| *Utensil storage* | No significant association | No significant association |
| *Water source* | No significant association | No significant association |
| *Water treatment* | No significant association | No significant association |
| *Toilet facility* | No significant association | No significant association |

**Source:** Field survey, 2025

## 4.0 DISCUSSION

This study identifies a severe and multidimensional malnutrition crisis in Irele and Okitipupa LGAs, Ondo State. The 93.3% household food insecurity rate, combined with child stunting (39.3%), wasting (29.1%), and maternal overweight/obesity (58.4%), confirms the presence of the nutrition transition’s “double burden” within the same communities. These findings align with Popkin et al. (2020), who argue that developing economies are increasingly characterized by simultaneous undernutrition and overnutrition driven by the consumption of cheap, energy-dense, nutrient-poor foods.

The finding that socioeconomic variables (income and education) were not significantly associated with child anthropometric outcomes is notable. It suggests that within this largely homogeneous low-income population, the environmental and structural determinants—including food system constraints and IYCF practices—exert a greater influence on nutritional status than individual socioeconomic factors. This is consistent with the environmental enteric dysfunction hypothesis (Humphrey et al., 2019), which proposes that a poor environment can override individual biological or socioeconomic protective factors.

The significant associations between wasting and breastfeeding duration (p = 0.008) and early introduction of family foods (p = 0.032) highlight specific, modifiable behavioral targets for intervention. Children of mothers who breastfed for shorter durations or who introduced family foods before six months were more likely to be wasted, supporting WHO recommendations on optimal breastfeeding and complementary feeding timing.

The cross-sectional design limits causal inference; associations identified should be interpreted as associational rather than causal. Self-reported dietary and IYCF data are subject to recall and social desirability bias—notably evidenced by the consistently high on-demand breastfeeding rates across all nutritional groups. The study did not incorporate biochemical indicators of micronutrient deficiency (e.g., serum ferritin, haemoglobin), limiting detection of hidden hunger. The relatively homogeneous socioeconomic profile of the sample may have reduced statistical power to detect income- or education-related differences. Additionally, seasonal variation in food availability was not captured in this single-timepoint design.

The findings must be interpreted with caution given the cross-sectional design and the limitations in self-reported data. However, they provide a strong foundation for LGA-specific public health planning. The dual burden of malnutrition observed—severe child undernutrition coexisting with maternal overweight—is consistent with trends documented across sub-Saharan Africa (Popkin et al., 2020; WHO, 2021) and reinforces the inadequacy of single-dimensional nutrition interventions. The non-significant relationship between socioeconomic status and child anthropometric outcomes, combined with the significant role of IYCF practices, shifts the policy emphasis toward behavioral and environmental interventions rather than income-focused approaches alone.

The study’s findings are generalizable to similar rural and semi-urban LGAs in southwestern Nigeria sharing comparable agrarian economies, infrastructure deficits, and food environment characteristics. The multistage sampling approach enhances representativeness within the study area. However, findings should be applied cautiously to peri-urban or northern Nigerian contexts, where distinct dietary staples, cultural practices, and economic structures differ significantly.

This study confirms that high household food insecurity and a dual burden of malnutrition coexist in rural Ondo State. Stunting (39.3%), wasting (29.1%), and underweight (42.1%) rates among children under five indicate a public health emergency, while 58.4% of mothers are overweight or obese—a pattern driven by the nutrition transition toward cheap, energy-dense but nutrient-poor diets. IYCF practices—specifically the duration of breastfeeding and timing of complementary food introduction—are significant modifiable determinants of child wasting, offering concrete targets for intervention. These findings dispute one-size-fits-all national nutrition strategies and demonstrate the need for decentralized, LGA-specific policies that simultaneously address food insecurity, dietary quality, and optimal infant feeding in rural communities.

Local governments and health agencies in Ondo State should strengthen community-based nutrition education programs focused on the first 1,000 days, emphasizing optimal breastfeeding duration and the appropriate timing of complementary food introduction. Policymakers should implement food subsidies for protein-rich food groups and support local agricultural incentives to address the 93.3% food insecurity rate. Community-driven home gardening and small-scale livestock programs can improve dietary diversity. Integration of routine nutrition screening and community-based management of acute malnutrition (CMAM) into primary healthcare services is urgently needed. Future research should adopt longitudinal designs, include biochemical indicators, and expand geographic coverage to other LGAs in Ondo State.

## Data Availability

All data produced in the present study are available upon reasonable request to the authors

## Statements and Declarations

### Author Contributions

Gbayisemore Erisan Eunice conceptualized the study, designed its framework, synthesized the literature, prepared the original draft, supervised data collection across both sites, and approved the final version for publication. Ajayi Kayode developed the sampling framework, performed formal statistical analyses, interpreted the data, and edited the manuscript for methodological rigor. Ibidayo Adefolake ALEBIOSU supported data interpretation, provided critical review, and ensured fidelity to research objectives. Abolaji Moses Ogunetimoju conducted final editing and polishing, coordinated journal submission, and confirmed compliance with publication guidelines. All authors reviewed and approved the final manuscript.

### Funding

This research was conducted without external funding or specific grants from public, commercial, or not-for-profit funding agencies. The study was self-funded by the researchers, who retained full control over study design, data collection, analysis, interpretation, and submission.

### Clinical trial number

Not applicable.

### Institutional Review Board Statement

The study was conducted in accordance with the Declaration of Helsinki and approved by the Health Research Ethics Committee (HREC) of the Ministry of Health, Ekiti State, Nigeria. Informed consent was obtained from all participants. Confidentiality and anonymity were strictly maintained throughout the study.

### Conflicts of Interest

The authors declare no conflict of interest.

## Acknowledgments

The authors wish to express their sincere gratitude to the participants from Irele and Okitipupa for their cooperation and willingness to share information. We acknowledge the field assistants for their dedication during data collection, and the community leaders and local authorities for facilitating community entry and the smooth conduct of the research. We appreciate the management and staff of the Department of Human Nutrition and Dietetics at Afe Babalola University, Ado Ekiti, and the Department of Statistics at Obafemi Awolowo University, Ile-Ife, for providing institutional support.

## Notes

### Competing Interest Statement

The authors have declared no competing interest.

### Funding Statement

This study did not receive any funding

### Author Declarations

Ethical approval was obtained from the Health Research Ethics Committee (HREC) of the Ministry of Health, Ekiti State, Nigeria, in accordance with the Declaration of Helsinki.

